# A regression discontinuity analysis of the social distancing recommendations for older adults in Sweden during COVID-19

**DOI:** 10.1101/2021.07.22.21260973

**Authors:** Carl Bonander, Debora Stranges, Johanna Gustavsson, Matilda Almgren, Malin Inghammar, Mahnaz Moghaddassi, Anton Nilsson, Paul W Franks, Maria Gomez, Tove Fall, Jonas Björk, COVID Symptom Study Sweden

**Author notes:** **Corresponding author. Address:** Carl Bonander, School of Public Health and Community Medicine, Institute of Medicine, Sahlgrenska Academy, University of Gothenburg, SE-405 30 Gothenburg, Sweden.

## Abstract

**Objectives:** To study the impact of non-mandatory, age-specific social distancing recommendations for older adults (70+ years) in Sweden on isolation behaviors and disease outcomes during the first wave of the COVID-19 pandemic.

**Methods:** Our study relies on self-reported isolation data from COVID Symptom Study Sweden (n = 96,053) and national register data on COVID-19 hospitalizations, deaths, and confirmed cases. We use a regression discontinuity design to account for confounding factors, exploiting the fact that exposure to the recommendation was a discontinuous function of age.

**Results:** By comparing individuals just above to those just below the age limit for the policy, our analyses revealed a sharp drop in the weekly number of visits to crowded places at the 70-year-threshold (−13%). Severe COVID-19 cases (hospitalizations or deaths) also dropped abruptly by 16% at the 70-year-threshold. Our data suggest that the age-specific recommendations prevented approximately 1,800 to 2,700 severe COVID-19 cases, depending on model specification.

**Conclusion:** The non-mandatory, age-specific recommendations helped control the COVID-19 pandemic in Sweden.

## Introduction

Non-pharmaceutical interventions (NPIs) represent a broad range of individual, environmental, and population-related public measures that aim to reduce the transmission of infectious diseases.^1^ When implemented at the population level, these measures intend to reduce the spread of infection by limiting the proximity of interpersonal interactions within or between population groups. During the first wave of the COVID-19 pandemic in spring 2020, 19 countries in the EU/EEA and the UK implemented specific ‘stay-at-home’ recommendations for risk groups or vulnerable populations.^2^ The Swedish Public Health Agency issued a non-mandatory recommendation for individuals aged 70 or over, i.e., the most vulnerable population group concerning severe COVID-19 disease, to avoid contact with persons outside the household and crowded places (e.g., stores, public transportation).^3^

It is difficult to predict to what degree the public will comply with recommendations that require substantial modifications in daily life. Compliance depends on many factors, including how the population perceives the risk and the consequences of isolation, the government’s credibility, and the clarity of communication.^4,5^ Most people are more willing to protect themselves if they belong to a risk group.^4^ However, older adults do not seem more inclined to self-isolate or comply with social distancing recommendations than people in their 50’s and 60’s, according to data from 27 countries collected during the first wave of the COVID-19 pandemic.^6^ The added effect of targeting risk communication at older adults also appears limited.^7^ These results raise questions regarding the effectiveness of Sweden’s non-mandatory isolation policy for older adults.

Surveys indicate that many followed the recommendation,^8,9^ and epidemiologic simulations suggest the policy might have reduced the number of infections and deaths,^9^ but empirical evidence is lacking. We address this gap by studying the impact of Sweden’s age-specific recommendation on social distancing behaviors and disease outcomes among older adults during the first wave of the COVID-19 pandemic. Age affects social distancing behaviors and COVID-19 disease risks, but individuals just above or below the 70-year-threshold should be comparable on medical risk factors and other confounders. Our study uses the discontinuous nature of the government’s message to self-isolate created by the age-specific recommendations. Using a regression discontinuity design,^10^ we aim to test if non-mandatory, age-specific recommendations can affect social distancing behaviors and disease outcomes among older adults.^11^

## Methods

### The recommendations and context

On March 16, 2020, the Public Health Agency in Sweden issued a special recommendation that individuals aged 70 years or older should avoid contact with persons outside the household and crowded places. The same recommendation was given to individuals younger than 70 years if they had at least one of the following risk factors: high blood pressure, heart disease, lung disease, obesity, diabetes, or receiving immunosuppressant treatment (e.g., cancer patients). This group-specific recommendation was only active during the first wave of the pandemic, which occurred between the middle of March and the end of July 2020.^9^

### Social distancing outcome measures

We used data from COVID Symptom Study Sweden (CSSS),^12^ an app-based study that collects data for epidemiologic surveillance and prediction of severe acute respiratory syndrome coronavirus 2 (SARS-CoV-2) infection via daily self-reports of disease symptoms.^13–15^ On their first use of the app, participants self-reported their year of birth, sex, height, weight, and geographical location (postal code). They also completed a health survey with questions about pre-existing health conditions.

As the first wave occurred during the spring of 2020, we considered individuals who were 70 years of age at the end of 2019 (i.e., born in 1949) to be exposed to the age-specific social distancing recommendations. From May 7 to September 29, 2020, the app also included a weekly question about the levels of isolation during the last seven days. The respondents were asked: (i)“In the last week, how many times have you visited somewhere with lots of people (e.g., groceries, public transport, work)?”, (ii)“In the last week, how many times have you been outside, with little interaction with people outside your household (e.g., exercise)?”, and (iii)“In the last week, how many times have you visited a healthcare provider (e.g., hospital, clinic, dentist, pharmacy)?”. As described further below, our analysis focuses on individuals close to the 70-year-threshold. However, it requires data from younger and older individuals to model the relationship between these social distancing measures and age. We decided *a priori* to include period-specific averages of the social distancing measures for individuals born before 1980 (i.e., age 39 at the end of 2019) in the study, and there were too few participants born each year before 1940 (79 years) to be included in the analysis of the social distancing data. Participants also had to have at least one observation of isolation data from the period when the social distancing questions were asked up until the end of the first wave of the pandemic ([May 7 – July 31 2020]) (*n* = 96,053). **Table 1** contains an overview of characteristics of the entire sample and for individuals close to the age threshold for the recommendations (65-69 years, 70-74 years).

**Table 1.** Characteristics of the sample born 1940 - 1980 and average social distancing behaviors stratified by risk group.

| Characteristic | *Total*<br>N=96,053 | *65-69 yrs*<br>N=9,358 | *70-74 yrs*<br>N=5,992 |
| --- | --- | --- | --- |
| ***Demographics*** |  |  |  |
| Age at the end of 2019 - mean (SD) | 54.5 (10.2) | 66.9 (1.4) | 71.4 (1.1) |
| Women - % | 60.2% | 55.7% | 50.5% |
| Lives in Stockholm County - % | 22.9% | 19.9% | 19.5% |
| ***Risk factors*** |  |  |  |
| Obese (body mass index $\geq 30$) - % | 19.4% | 16.2% | 13.7% |
| Diabetes - % | 4.6% | 8.8% | 8.6% |
| Lung disease - % | 13.8% | 13.5% | 12.7% |
| Cancer - % | 1.4% | 3.0% | 3.6% |
| Heart disease - % | 6.4% | 12.2% | 15.2% |
| Takes immunosuppressants - % | 4.7% | 5.8% | 6.0% |
| Has at least one risk factor - % | 38.0% | 42.5% | 42.0% |
| ***Level of isolation, by type of activity*** |  |  |  |
| Visited crowded places, n times (weekly) - mean (SD) | 5.4 (6.3) | 4.3 (6.6) | 2.9 (5.2) |
| Visited crowded places, n times (weekly) - median (IQR) | 4.0 (2.3-6.3) | 2.8 (1.6-4.8) | 1.9 (0.8-3.3) |
| Outdoors with limited interaction, n times (weekly) - mean (SD) | 8.8 (9.6) | 9.8 (11.6) | 10.0 (12.0) |
| Outdoors with limited interaction, n times (weekly) - median (IQR) | 6.3 (4.0-10.0) | 6.6 (4.2-10.5) | 6.5 (4.0-11.0) |
| Visited healthcare provider, n times (weekly) - mean (SD) | 0.5 (1.1) | 0.4 (0.7) | 0.3 (0.4) |
| Visited healthcare provider, n times (weekly) - median (IQR) | 0.2 (0.0-0.6) | 0.2 (0.0-0.5) | 0.1 (0.0-0.4) |

*Notes:* The level of isolation reflects weekly average of available self-reports between the period 2020-05-07 and 2020-07-31.

We averaged each of the three social distancing measures within each respondent to a weekly average during the follow-up period above. Because we mainly expected the isolation policy to affect visits to crowded places, we viewed measure (i) as our primary social distancing outcome. According to the recommendations, going outdoors with limited physical interaction was fine. As a result, measure (ii) should not be affected. It was less clear what to expect for measure (iii). It was recommended that a courier (such as a younger relative) collect prescriptions from pharmacies. However, measure (iii) also included in-person healthcare visits, for which postponement can be considered an unintended consequence of the intervention.

### Disease outcomes

We also investigated population-level effects on severe cases (hospitalizations or deaths attributable to COVID-19). We obtained national data on all individuals born before 1980 and coded a binary indicator for whether they had at least one inpatient COVID-19 disease episode or had died due to COVID-19 disease during the first wave (March 16 – July 31, 2020; *n* population = 5,396,837; *n* severe cases = 21,804). The inpatient data were retrieved from the National Patient Register^16^ and mortality data from the Cause of Death Register^17^ (see Supplementary material for a detailed description of these data). The retrieved data also contained information on year of birth, home address postal code, and sex.

As a secondary disease outcome, we used the number of confirmed infections by polymerase chain reaction (PCR) testing obtained from the SmiNet database at the Public Health Agency (*n* confirmed cases = 48,984). It was mandatory for all clinical laboratories in Sweden to report PCR tests positive for SARS-CoV-2 to SmiNet during the COVID-19 pandemic.^12^ Testing, however, was highly selective during the first wave, and positive cases mainly represented those who either needed treatment or were being tested due to their work in the health care industry. Thus, absolute effects should be interpreted with caution. Nevertheless, as explained in the following section, our design compares individuals aged just above and below 70 years. Therefore, relative estimates can still be meaningful, assuming that testing probabilities were equal close to this threshold.

### Regression discontinuity analysis

We relied on a sharp regression discontinuity design (RDD) to estimate the effect of the recommendations on social distancing behaviors and disease outcomes as a discontinuous function of age (in years) at the 70-year-threshold. The design has, for instance, previously been used to estimate the effects of early versus deferred antiretroviral therapy for HIV patients,^18^ and the impact of age-specific policies (e.g., minimum drinking age laws and co-payments in healthcare).^19,20^

By exploiting abrupt changes induced by arbitrary thresholds, such as an age limit, causal effects can be estimated in observational data without controlling for confounders.^18^ RDD estimates reflect local average effects at the threshold (i.e., for people who are 70 years old exactly). The key causal assumption is that nothing else that affects the outcomes changes discontinuously at the policy threshold other than the social distancing recommendation.^11^ We are not aware of any other policies that might have affected social distancing or COVID-19 disease at the 70-year-threshold. We note that retirement may affect the tendency to self-isolate, which would be a major confounder in a typical observational study. The retirement age in Sweden is flexible, but the most common is 65. As the probability of retiring does not change abruptly at 70 years of age, the RDD estimates will not be biased by retirement decisions.

Our implementation follows the RDD estimation and reporting guidelines outlined by Athey & Imbens^21^, Boon et al^10^, and Gelman & Imbens^22^. While our primary interest is in individuals just above and below the 70-year-threshold, RDD estimation requires fitting models to estimate the relationship between outcome variables and age. This estimation is usually performed within a small age window around the threshold (also known as *bandwidth*). The outcome-age relationship is not of primary interest but helps capture the effects of confounding variables that develop smoothly with age. However, it is crucial to use an appropriate model specification and bandwidth to avoid bias due to model misspecification.^22^ Complex model specifications in RDD analyses are prone to overfitting, and Gelman & Imbens caution against using models with high-order polynomials (greater than linear or quadratic).^22^ We therefore used local linear and quadratic regressions to estimate the jump in the outcomes at the threshold. In each analysis, we used a data-driven bandwidth selection method to identify the mean squared error (MSE) optimal window around the 70-year-threshold.^23^ The larger the bandwidth (i.e., the age window used in the analysis), the more individuals are included, which increases the precision of the effect estimates. However, the risk of bias due to model misspecification also increases. The data-driven procedure aims to identify the largest possible window in which the relationship between the outcome and age is approximately linear (or quadratic, depending on the model). The analyses were performed using the *rdrobust* package (version: winter 2020) for Stata (version 16.1).^24^ The program provides additive effect estimates by default. We also converted these estimates to relative effects to aid the interpretation of the additive estimates. More details about the estimation strategy are provided in the Supplementary material.

#### Subgroup analyses

Men are at higher risk of severe COVID-19 disease, which may positively influence their willingness to comply with the recommendations.^4^ Similarly, Stockholm experienced the most severe outbreak during the first pandemic wave in Sweden (7.66 severe cases per 1000 population in Stockholm in the age range 65-74 years versus 3.02 per 1000 population in the rest of Sweden according to our data). We therefore conducted subgroup analyses by sex and geographical area to assess differences in effect size.

In the social distancing data, which contained information on medical risk factors, we also considered two additional subgroups: those without any and those with at least one of the following six risk factors communicated by the Public Health Agency in May 2020: obesity (body mass index ≥ 30), diabetes, lung disease, cancer, heart disease, or on immunosuppressant medication. Because the recommendation also covered younger individuals with other risk factors, we expected the age-specific recommendations to have a greater impact on people with no risk factors except old age.

#### Sensitivity analyses

We performed recommended sensitivity, balance, and falsification checks to assess the risk of bias.^10^ We present these analyses in the Supplementary material. In summary, the results from analyses with alternative bandwidths are similar to the main results. The data also passed standard falsification and balance checks (e.g., no evidence of sorting around the threshold, no clear evidence of discontinuities on covariates).

### Ethical approval

The Swedish Ethical Review Authority has approved CSSS (DNR 2020-01803 with addendums 2020-04006, 2020-04145, 2020-04451, and 2020-07080).

## Results

### Principal findings

The study participants went to crowded places 5.4 times a week, outdoors with limited interaction 8.8 times a week, and to healthcare providers 0.5 times a week on average during follow-up (see **Table 1** for details). **Figure 1** shows how these behaviors varied by age, alongside the fitted values from local linear (**Figure 1A**) and quadratic (**Figure 1B**) regressions estimated within the optimal windows around the 70-year-threshold. The analysis suggests that the policy threshold is associated with a sharp decline in the average number of times older adults visited crowded places (e.g., stores) during the first wave of the pandemic (−0.47 [95% CI: -.89, -.05] times less per week in the entire sample, which corresponds to a 13% reduction; **Table 2**; **Figure 1**). We found no evidence of discontinuities at the 70-year-threshold on being outside with little interaction or visits to healthcare providers (**Table 2**; **Figure 1**).

**Figure 1.**
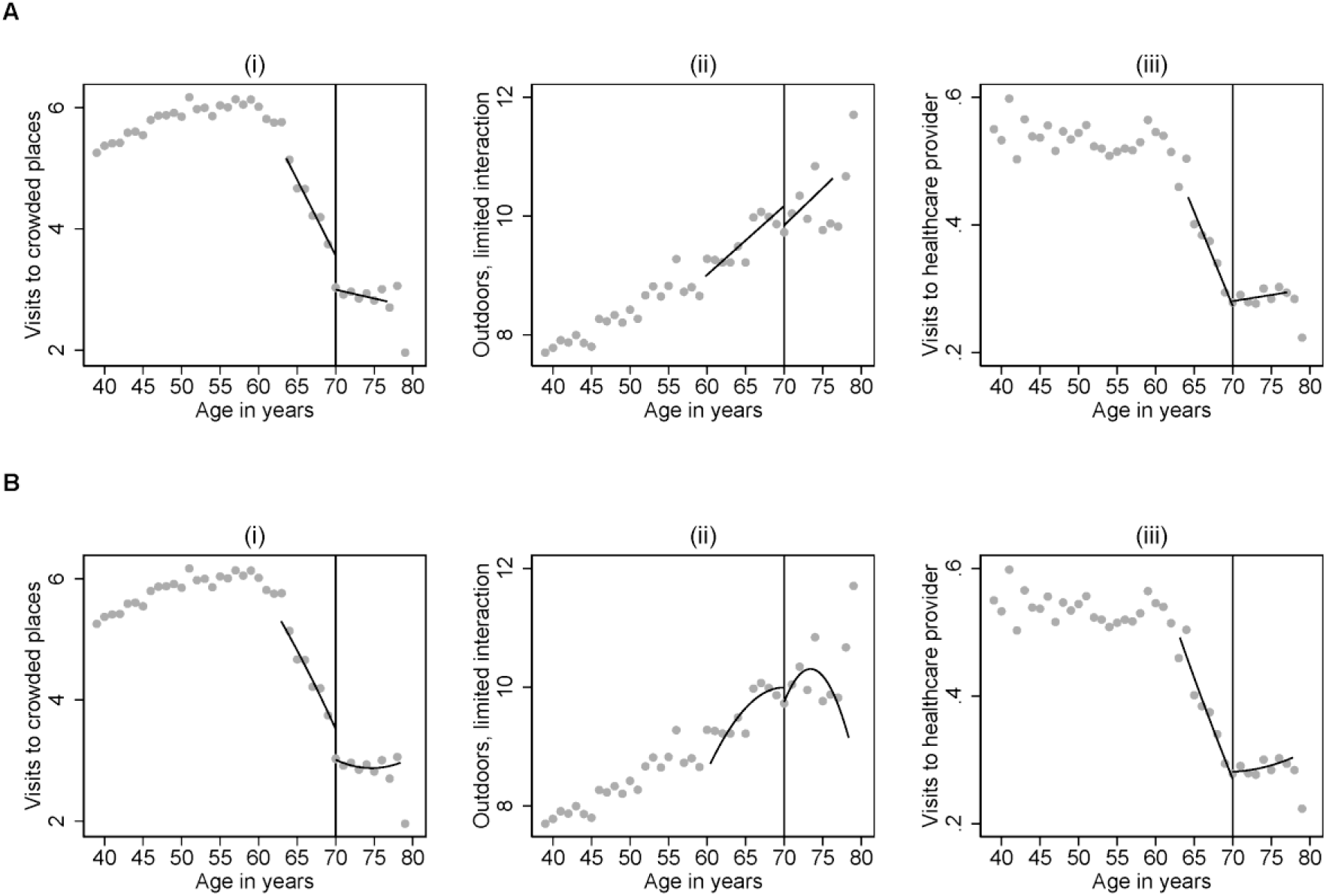
Regression discontinuity plots for the impact of Sweden’s age-specific isolation recommendations on social distancing behaviors at the 70-year-threshold with local linear (panel A) and quadratic (panel B) estimates in mean-squared-error-optimal bandwidths around the threshold, for three social distancing measures: (i) mean weekly visits to crowded places, (ii) mean weekly outdoor episodes with no or limited interaction, and (iii) mean weekly visits to healthcare providers.

**Table 2.** Regression discontinuity estimates of the effect of the social distancing recommendations for people aged 70+ years in Sweden on the level of isolation during the first wave of the COVID-19 pandemic, by type of activity, model specification and subgroup.

| Group | Model specification |  |  |  |
| --- | --- | --- | --- | --- |
|  | Local linear |  | Local quadratic |  |
|  | Additive effect | Relative effect | Additive effect | Relative effect |
| *i. Weekly visits to crowded places (e.g., stores, public transportation)* |  |  |  |  |
| Full sample | -.47<br>(-.89, -.05) | .87<br>(.77, .98) | -.57<br>(-1.25, .11) | .84<br>(.71, 1.04) |
| **Subgroup** |  |  |  |  |
| No risk factors | -.57<br>(-1.12, -.02) | .84<br>(.73, .99) | -.73<br>(-1.54, .09) | .81<br>(.67, 1.03) |
| At least one risk factor | -.32<br>(-.95, .31) | .90<br>(.76, 1.12) | -.56<br>(-1.50, .37) | .84<br>(.66, 1.14) |
| Men | -.56<br>(-1.14, .02) | .86<br>(.74, 1.01) | -.72<br>(-1.47, .04) | .82<br>(.69, 1.01) |
| Women | -.35<br>(-.91, .22) | .89<br>(.75, 1.09) | -.58<br>(-1.45, .28) | .83<br>(.66, 1.11) |
| Stockholm County | -.83<br>(-1.74, .08) | .79<br>(.64, 1.03) | -.72<br>(-1.74, .31) | .81<br>(.63, 1.11) |
| Rest of Sweden | -.38<br>(-.83, .08) | .89<br>(.79, 1.03) | -.60<br>(-1.37, .17) | .84<br>(.69, 1.06) |
| *ii. Weekly number of times gone outside with limited interaction* |  |  |  |  |
| Full sample | -.50<br>(-1.22, .21) | .95<br>(.89, 1.02) | -.16<br>(-1.21, .89) | .98<br>(.89, 1.10) |
| **Subgroup** |  |  |  |  |
| No risk factors | -.62<br>(-1.52, .29) | .94<br>(.87, 1.03) | .21<br>(-1.22, 1.64) | 1.02<br>(.89, 1.2) |
| At least one risk factor | -.41<br>(-1.68, .86) | .96<br>(.85, 1.10) | -.40<br>(-1.79, .98) | .96<br>(.84, 1.11) |
| Men | -.85<br>(-2.05, .35) | .92<br>(.82, 1.04) | -1.02<br>(-2.39, .35) | .90<br>(.80, 1.04) |
| Women | -.42<br>(-1.45, .62) | .96<br>(.87, 1.07) | 1.05<br>(-.58, 2.69) | 1.12<br>(.94, 1.38) |
| Stockholm County | -1.51<br>(-3.74, .71) | .87<br>(.73, 1.08) | -1.34<br>(-3.71, 1.02) | .88<br>(.73, 1.11) |
| Rest of Sweden | -.24<br>(-1.03, .55) | .98<br>(.9, 1.06) | -.04<br>(-1.11, 1.03) | 1.00<br>(.90, 1.12) |
| *iii. Weekly visits to healthcare provider(s)* |  |  |  |  |
| Full sample | .01<br>(-.02, .05) | 1.05<br>(.92, 1.21) | .01<br>(-.06, .07) | 1.03<br>(.83, 1.35) |
| **Subgroup** |  |  |  |  |
| No risk factors | .01<br>(-.04, .06) | 1.04<br>(.86, 1.30) | .01<br>(-.07, .08) | 1.02<br>(.78, 1.49) |
| At least one risk factor | .02<br>(-.03, .08) | 1.07<br>(.90, 1.30) | .03<br>(-.04, .10) | 1.10<br>(.88, 1.47) |
| Men | -.02<br>(-.07, .02) | .91<br>(.79, 1.09) | -.03<br>(-.08, .02) | .90<br>(.77, 1.09) |
| Women | .05<br>(-.01, .10) | 1.20<br>(.98, 1.53) | .02<br>(-.05, .10) | 1.08<br>(.86, 1.47) |
| Stockholm County | .03<br>(-.04, .10) | 1.13<br>(.89, 1.53) | .05<br>(-.06, .15) | 1.18<br>(.83, 2.04) |
| Rest of Sweden | .01<br>(-.04, .05) | 1.02<br>(.89, 1.21) | .00<br>(-.06, .06) | 1.00<br>(.83, 1.27) |

Notes: Additive estimates reflect bias-corrected effects on the difference scale (where 0 = null effect) estimated within MSE-optimal bandwidths, with 95% robust confidence intervals from the *rdrobust* package for Stata in parentheses. Relative estimates reflect ratios (where 1 = null effect) computed using the additive estimates (see Supplementary material for details).

Figure 2. shows estimated effects on the incidence of severe COVID-19 disease and all confirmed cases per 1,000 population at the national level during the first wave of the pandemic, and **Table 3** contains the effect estimates expressed as incidence rate differences and rate ratios. Overall, it appears that the recommendations may lead to a reduction in COVID-19 disease at the age threshold compared to a scenario without the age-specific recommendations (**Table 3; Figure 2**). The local linear estimates indicate a 16% reduction in both severe COVID-19 cases (incidence rate ratio [IRR] = 0.84 [95% CI: 0.73, 1.00]) and the number of confirmed cases (IRR = 0.84 [95% CI: 0.69, 1.08]) at the 70-year-threshold, although the confidence interval for confirmed cases overlaps the null (**Table 3**). For severe cases, the estimate was slightly larger in the quadratic specification (IRR = 0.78 [95% CI: 0.64, 0.99]). Our calculation in **Box 1** uses these numbers to estimate the impact of the recommendations, assuming that the relative effect is the same for everyone older than 70. The results imply that the policy prevented 1803 [95% CI: 19, 3636] severe cases (2737 [95% CI: 87, 5388] according to the quadratic estimate) and 624 [95% CI: 7, 1257] deaths (quadratic estimate: 947 [95% CI: 30, 1864]).

**Table 3.** Regression discontinuity estimates of the effect of the social distancing recommendations for people aged 70+ years in Sweden on severe COVID-19 disease and confirmed cases during the first wave of the COVID-19 pandemic, by model specification and subgroup.

| Group | Model specification |  |  |  |
| --- | --- | --- | --- | --- |
|  | Local linear |  | Local quadratic |  |
|  | IRD<br>(Absolute effect) | IRR<br>(Relative effect) | IRD<br>(Absolute effect) | IRR<br>(Relative effect) |
| *i. Severe cases (hospitalized or dead) per 1,000 population* |  |  |  |  |
| Full sample | -.65<br>(-1.29, -.01) | .84<br>(.73, 1.00) | -.99<br>(-1.94, -.03) | .78<br>(.64, .99) |
| **Subgroup** |  |  |  |  |
| Men | -.84<br>(-1.73, .05) | .84<br>(.73, 1.01) | -1.47<br>(-2.86, -.08) | .76<br>(.61, .98) |
| Women | .03<br>(-.58, .64) | 1.01<br>(.80, 1.38) | -.32<br>(-1.32, .67) | .88<br>(.64, 1.40) |
| Stockholm County | .52<br>(-1.02, 2.06) | 1.08<br>(.87, 1.40) | -.44<br>(-2.74, 1.86) | .943<br>(.72, 1.35) |
| Rest of Sweden | -.86<br>(-1.52, -.21) | .76<br>(.64, .93) | -1.24<br>(-2.20, -.29) | .69<br>(.55, .90) |
| *ii. Confirmed cases per 1,000 population* |  |  |  |  |
| Full sample | -.81<br>(-1.93, .31) | .84<br>(.69, 1.08) | -.73<br>(-2.02, .56) | .86<br>(.68, 1.15) |
| **Subgroup** |  |  |  |  |
| Men | -.67<br>(-1.62, .28) | .89<br>(.77, 1.05) | -1.46<br>(-3.11, .20) | .79<br>(.64, 1.04) |
| Women | -.44<br>(-1.73, .85) | .88<br>(.66, 1.35) | -.43<br>(-1.97, 1.12) | .89<br>(.63, 1.52) |
| Stockholm County | .83<br>(-.86, 2.52) | 1.12<br>(.90, 1.46) | -.29<br>(-3.34, 2.76) | .96<br>(.70, 1.53) |
| Rest of Sweden | -.78<br>(-2.02, .46) | .83<br>(.65, 1.14) | -.28<br>(-1.86, 1.29) | .91<br>(.60, 1.84) |

Notes: Estimates reflect bias-corrected incidence rate differences per 1,000 population (IRD, i.e., absolute effects where 0 = null effect) and incidence rate ratios (IRR) (i.e., relative effects where 1 = null effect) estimated within MSE-optimal bandwidths, with 95% robust confidence intervals from the *rdrobust* package for Stata in parentheses.

**Figure 2.**
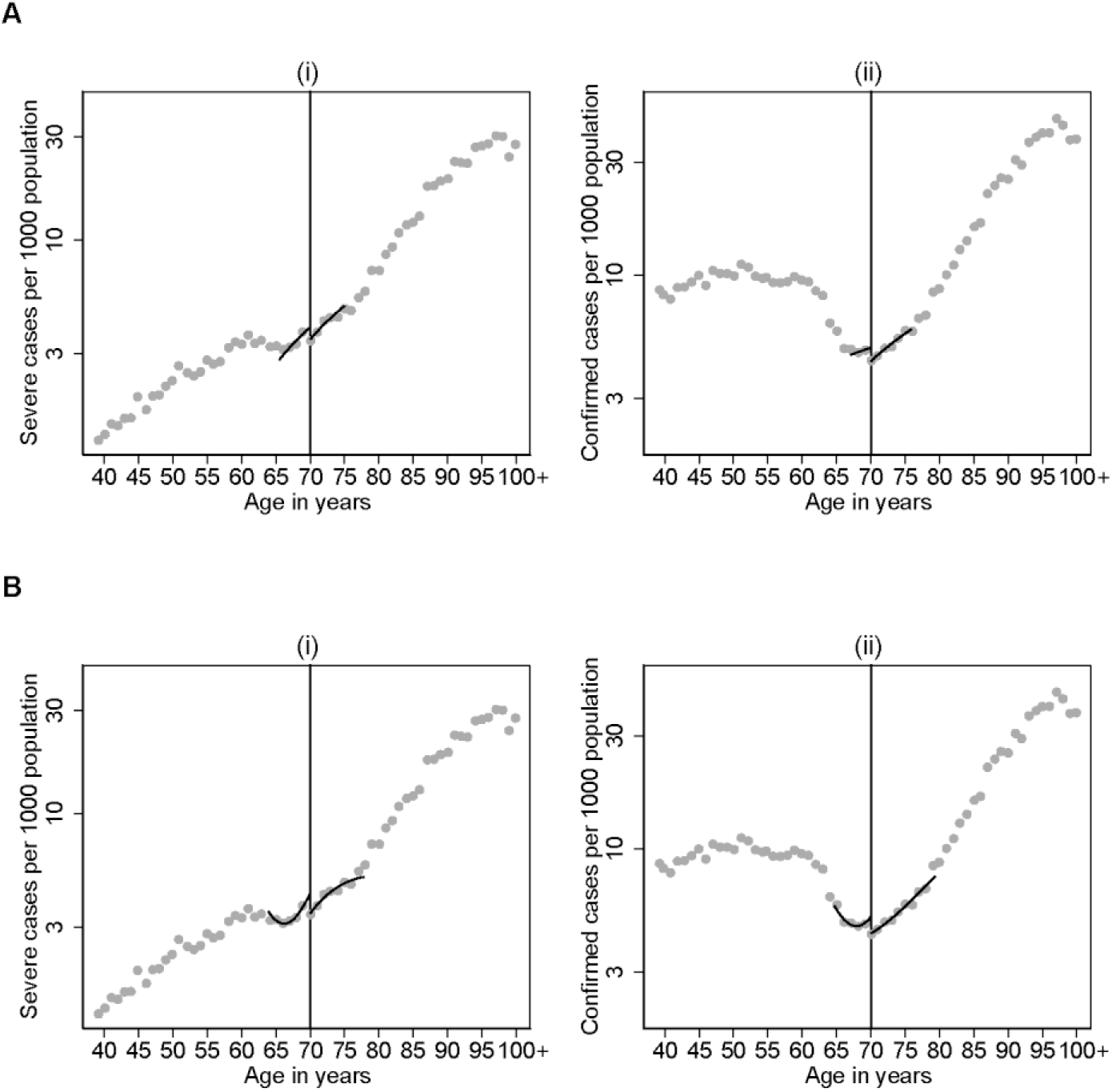
Regression discontinuity plots for the impact of Sweden’s age-specific isolation recommendations on COVID-19 disease incidence per 1,000 population at the 70-year-threshold with local linear (panel A) and quadratic (panel B) estimates in mean-squared-error-optimal bandwidths around the threshold, for two disease outcome measure: (i) severe cases (hospitalized or dead), and (ii) all confirmed cases. The y-axis is plotted on a logarithmic scale to enable better visualization of the data close to the 70-year-threshold.

### Subgroup analyses

The results from the subgroup analyses are presented alongside the main results in **Table 2** (social distancing outcomes) and **Table 3** (disease outcomes). Regression discontinuity plots for the subgroup analyses can be found in **Figures S1-S5**. We note that the estimates in the subgroup analyses were generally imprecise, and the observed differences between subgroups should therefore be interpreted with due caution.

Nonetheless, the point estimates imply that the effect on visits to crowded places was larger among individuals without risk factors than those with at least one risk factor, larger among men than women, and larger in Stockholm County than in the rest of Sweden (**Table 2; Figures S1-S2 in the supplementary material**). In an analysis exploring these subgroups further, we found evidence of an effect on visits to crowded places only among men (independent of risk group status) and women without other risk factors, but not among women with other risk factors (**Figure S3**). It is also worth noting that women with risk factors isolated themselves more than other groups, even at younger ages (**Figure S3)**.

For the disease outcomes, stronger absolute effects were suggested among men than among women (**Table 2; Figures S4-S5**), which is consistent with the social distancing results. However, no effect on COVID-19 disease was observed for Stockholm County, where results were inconclusive (**Table 3, Figures S4-S5**).

## Discussion

The results suggest that Swedish 70-year-olds isolated themselves more than those just below 70 years, implying that at least parts of the population adhered to the non-mandatory, age-specific recommendations communicated by the Swedish Public Health Agency.

The results were generally in line with expectations. In particular, we found that the effect was limited to visits to crowded places, which is the social distancing outcome we assumed would be affected most by the recommendations. The impact on social distancing also seems to have caused a drop in disease outcomes at the 70-year-threshold. We were unable to draw firm conclusions from our subgroup analyses, however. Results were inconsistent and inconclusive for Stockholm County, where the pandemic hit particularly hard during the first wave in Sweden. We found an indication that men may have been more affected by the age-specific recommendation than women, which is consistent with assumptions that high-risk groups are generally more willing to follow recommendations on protecting themselves.^4^ People with other risk factors (especially among women) also appeared to be more willing to self-isolate even at younger ages, which could – at least in part – be a consequence of the recommendations aimed at people with other risk factors.

Our study adds to the body of knowledge about the effectiveness of non-pharmaceutical interventions for the control of novel viruses. Previous evidence regarding the effectiveness of general stay-at-home recommendations and public information campaigns on disease transmission is mostly inconsistent and inconclusive.^25^ We are not aware of any empirical studies evaluating the effects of more specific restrictions or recommendations. A modeling study conducted by the Swedish Public Health Agency estimated that the age-specific recommendation prevented between 2,100 and 3,600 hospitalizations and 750 to 1,312 deaths during March – September 2020.^9^ Their study is based on assumptions about the reduction in the number of contacts. Our study provides direct empirical support that the recommendations helped control the outbreak, with impact estimates that are slightly smaller but close to the simulation study results (**Box 1**).

### Possible mechanisms and implications

The Swedish response to the COVID-19 pandemic was relatively lenient compared to most countries and mainly included non-mandatory recommendations to the public during the first wave of the pandemic.^26^ Part of the strategy was to shield vulnerable population groups while keeping society as open as possible. The age-specific recommendation was an important aspect of this strategy, and it is conceivable that the effects are dependent not only on the acceptance among those targeted but also on which other population-level measures (such as limiting the size of gatherings and restrictions directed towards non-essential businesses) that were implemented during the same period to limit disease transmission.^25^ The Swedish public also has high levels of social trust and trust in its government,^27–29^ which may have played an essential role in the success of the age-specific recommendations.^30^

The results also need to be interpreted in the light of concerns about the adverse effects of social isolation on mental health.^8,31–35^ The age-specific recommendations were withdrawn in October 2020 as a result of these concerns.^9^ However, the body of evidence on the pandemic’s impact on older adults’ mental health is inconsistent, according to a recent review.^36^ Nonetheless, it is still important to account for adverse effects and other unintended consequences to understand the recommendation’s net impact on health and welfare. When the age-specific recommendations were withdrawn, they were followed by recommendations to meet in“social bubbles”, i.e., limiting one’s social contacts by only meeting with the same people, whether family, friends, or co-workers.^1,37^ This approach to mitigate the adverse effects of social isolation has been recommended by authorities in several countries during the pandemic. However, its consequences have so far only been evaluated in simulation studies.^38^

### Strengths and limitations

Our study relied on an RD design, which allows for causal effect estimation in observational data under relatively weak assumptions.^10^ Other policies that use the same threshold may, however, bias the results.^11^ Sweden had no other relevant policies using a 70-year threshold during the COVID-19 pandemic. The observed discontinuities were also isolated to the expected outcome variables, suggesting causality. The validity of our estimates also depends on appropriate modeling of the age-outcome relationship. We followed the current best practice recommendations, which is to fit simple models (linear or quadratic) within a data-driven bandwidth (age window) around the threshold.^21,22^ In this modeling framework, the typical concern is that the conclusions may depend heavily on the selected bandwidth.^10^ The Supplementary material shows that the main results are robust to other reasonable bandwidth choices. A limitation is that RDD can only be used to estimate effects for persons who are exactly 70 years old. The estimates may not generalize to older parts of the targeted age group, and the calculations in **Box 1** should, therefore, be interpreted with caution.

A key strength of our study was the availability of detailed and complete register data for severe COVID-19 disease, which most likely limited the extent of outcome misclassification, together with repeated assessment of social distancing during the study period. However, participants in the app study were more healthy than the general population and less likely to be smokers,^12^ which may hamper the generalizability of the observed effect on social distancing. Moreover, the general user of the COVID Symptom Study app might be more likely to follow recommendations than the average Swedish citizen, which may lead to an overestimation of the effect in the social distancing data, but this does not affect the disease outcome results. Another limitation was that the social distancing data was only available after May 6, 2020, and thus presents the latter part of the first pandemic wave. The study was further limited by the selected PCR testing in Sweden during the spring of 2020, which meant that we could not quantify effects on infection rates in absolute terms. In addition, we only had access to data on year of birth, which means that there is likely some degree of exposure misclassification bias in our estimates. We also lacked data on cohabitation, which may have also led to exposure misclassification as it is more likely that someone who lives with a partner belonging to a risk group adheres to the recommendations than someone of the same age who is living alone. In both cases, we believe that the misclassification would lead to an underestimation of the true effect. Another limitation was that we could only stratify effects by medical risk factors for social distancing outcomes and not for disease outcomes, as such register data were not available for the present study.

## Conclusion

The age-specific social distancing recommendations appear to have successfully reduced the spread of SARS-CoV-2 in Sweden, suggesting that non-mandatory social distancing recommendations targeting risk groups may reduce disease transmission during a pandemic, protect against severe disease, and save lives.

## Supporting information

Suppmentary material

## Data Availability

Data can be made available for further research on request, but usually requires an addendum of the ethical approval as well as consent from the data providers.

## Acknowledgments

We are grateful to the participants of the COVID Symptom Study. We also thank Martin Adiels (University of Gothenburg) and colleagues for compiling and sharing their data on the share of severe cases that occurred in care homes. The group authorship for this study (COVID Symptom Study Sweden, CSSS) consists of the following members of the analytical team at CSSS: Beatrice Kennedy (Uppsala University [UU]), Hugo Fitipaldi (Lund University [LU]), Ulf Hammar (UU), Marlena Maziarz (LU), Neli Tsereteli (LU), Nikolay Oskolkov (SciLife/LU), Georgios Varotsis (UU), Lampros Spiliopoulos (LU).

## Contribution statement

**Carl Bonander:** Conceptualization; Data curation; Formal analysis; Funding acquisition; Methodology; Visualization; Project management; Writing – original draft preparation. **Debora Stranges:** Conceptualization; Writing – review & editing. **Johanna Gustavsson:** Investigation; Writing – review & editing. **Matilda Almgren & Malin Inghammar:** Writing – review & editing. **Mahnaz Moghaddassi:** Data curation; Writing – review & editing. **Anton Nilsson:** Methodology; Writing – review & editing. **Paul Franks, Maria Gomez & Tove Fall**: Funding acquisition; Investigation; Resources; Methodology; Writing – review & editing. **Jonas Björk:** Conceptualization; Methodology; Funding acquisition; Supervision; Project management; Writing – original draft preparation; Writing – review & editing. **COVID Symptom Study Sweden (Group authorship):** Data curation; Software; Investigation.

## Tables and figures

### Box 1

**Approximate calculation of the total impact of the age-specific recommendations on COVID-19 disease in the age group 70+ years during the first pandemic wave in Sweden.**

We here estimate the total impact of the recommendations on severe COVID-19 disease and deaths, assuming that the relative effect is constant for all ages above 70 years. During the study period, 21,804 cases of severe COVID-19 disease occurred in Sweden according to our definition (hospitalized or dead); 12,258 (56%) of these cases were aged 70+ years, 46% of whom died due to the disease. An ongoing study that links COVID-19 case data with data on living conditions indicate that 22.2% of severe cases in this age group occurred in care homes (personal communication, Martin Adiels, University of Gothenburg, 2021-06-22). It is unlikely that the recommendations could have prevented these cases. Combining these numbers with ours suggest that approximately 12258*(1-0.222)=9537 severe cases occurred in the community-dwelling population. Dividing this number by the relative local linear estimate (IRR = 0.841 [95% CI: 0.724, 0.998]) implies that 11340 severe cases would have occurred without recommendations. This suggests that the policy prevented 1803 [95% CI: 19, 3636] severe cases during the first wave (2737 [95% CI: 87, 5388] according to the quadratic estimate). Turning to the number of deaths, the external data show that a larger share of deaths (41.5%) than severe non-fatal cases (7.26%) occurred in care homes. Repeating the above calculations for the number of deaths in our data (n = 5,639), our policy effect estimates imply that 624 [95% CI: 7, 1257] deaths were prevented (quadratic estimate: 947 [95% CI: 30, 1864]).

