## Supplementary material for "A regression discontinuity analysis of the social distancing recommendations for older adults in Sweden during COVID-19": Suppmentary material

for

### Supplementary methods

#### Classification of severe COVID-19 cases

We extracted our primary disease outcome data from the Swedish National Patient Register[1] and the Cause of Death Register[2], both of which cover the entire Swedish population. Our primary endpoint was hospitalization (an inpatient episode) or death due to COVID-19 during the period March 16, 2020 to July 31, 2020. Classification was made according to the International Classification of Diseases and Related Health Problems, tenth revision (ICD-10). The inpatient data were retrieved from the National Patient Register, using the emergency ICD-10 codes U07.1 (COVID-19, confirmed by laboratory testing) and U07.2 (COVID-19, virus not identified). Mortality data were retrieved from the Cause of Death Register, where COVID-19 deaths are defined using ICD-10 codes U07.1, U07.2, and B34.2 (Coronavirus infection, unspecified). Persons diagnosed with U07.1 and/or U07.2 within 30 days of their date of death are also included in the definition of COVID-19 deaths, regardless of the cause(s) listed in the cause-of-death certificate. The databases were linked at the individual level by the National Board of Health and Welfare using personal identification numbers (PIN). The final dataset was cleaned of PINs and pseudonymized to before being sent to us.

#### Detailed description of the estimation strategy

We used a data-driven estimation strategy to determine the mean-squared-error (MSE) optimal bandwidth to include in each analysis. Specifically, we followed the approach suggested by Calonico, Cattaneo & Titiunik,[3] which determines an MSE-optimal window for each analysis depending on the model specification and data. Their estimator also includes a bias-correction to account for bias owing to the bias-variance trade-off in the MSE-optimal bandwidth, where the bias is estimated and accounted for by fitting a polynomial regression of one degree higher than the main model (e.g., a quadratic bias model for estimates based on local linear regression). This procedure allows for valid and robust statistical inference with data-driven bandwidth selection.[3] As recommended, we used a triangular kernel to linearly down-weight observations away from the policy threshold.[4] We also allowed the slopes and optimal bandwidth lengths to differ on either side of the threshold. The analyses were performed using the *rdrobust* package (version: winter 2020) for Stata (version 16.1).[5]

#### Method for calculating relative effects from RD estimates

The *rdrobust* package provides estimates of the additive effect on the outcome. Using the potential outcomes framework, the estimated quantity can be written as

(1) $\beta = E[Y_i(1)|Age_i = 70] - E[Y_i(0)|Age_i = 70]$,

where $Y_i(1)$ and $Y_i(0)$ are the potential outcomes if individual *i* is exposed and not exposed to the recommendations, respectively. A relative (ratio) version of this estimate can be written as:

(2) $E[Y_i(1)|Age_i = 70]/E[Y_i(0)|Age_i = 70]$

The conditional average of the realized outcomes $Y_i$ among individuals aged exactly 70 years provides a direct estimate of $E[Y_i(1)|Age_i = 70]$ because all individuals aged 70 years are exposed to the recommendations, but $E[Y_i(0)|Age_i = 70]$ needs to be inferred. Re-arranging Equation (1) yields the following expression:

(3) $E[Y_i(0)|Age_i = 70] = E[Y_i(1)|Age_i = 70] - \beta$

To estimate relative effects, we can therefore use

(4) $\hat{E}[Y_i|Age_i = 70]/(\hat{E}[Y_i|Age_i = 70] - \hat{\beta})$,

where $\hat{E}[Y_i|Age_i = 70]$ is the average outcome among individuals aged exactly 70 years, and $\hat{\beta}$ is the additive effect estimate from the regression discontinuity analysis. To get 95% confidence intervals, we replace $\hat{\beta}$ with the lower and upper bounds of the confidence intervals for $\hat{\beta}$.

### Supplementary results

#### Sensitivity analyses

In this section, we present the results from a broad range of sensitivity and falsification checks typical for RD designs.[6] Specifically, we (1) checked for discontinuities in the other covariates presented in **Table 1** to assess violations of the continuity assumption, (2) conducted covariate-adjusted regression discontinuity analyses to adjust for any jumps in covariates at the age threshold, (3) varied the age window by ±1 and ±2 years from the MSE-optimal bandwidths to assess sensitivity to stochastic errors in the determination of the optimal bandwidth, and (4) checked for sorting in the number of observations just above and below the threshold in the social distancing data to rule out effects of the recommendations on selection into the study sample using the *rddensity* package for Stata.[7,8]

For the social distancing outcomes, varying the bandwidth by ±1 and ±2 years around the MSE-optimal age window gave rise to similar estimates as in the main analysis for visits to crowded places (**Tables S1-S2 in the supplementary material**). Estimates of the effect on the other two social distancing outcomes changed sign in a few analyses and subgroups but were generally consistent with the main analyses in that they do not provide any strong indications of an effect (**Tables S1-S2**). The covariate falsification checks did not show any meaningful and robust jumps in covariates (**Table S3)**, and covariate-adjusted estimates were very similar to the main analyses (**Table S3**). Finally, we found no evidence of sorting of observations around the 70-year-threshold (**Figure S6**; p-value for a jump in the density of observations at the threshold: 0.95).

The disease outcome results appeared slightly more sensitive to bandwidth selection than the social distancing estimates (especially to increasing the age window), which was not surprising given that the disease outcomes are highly non-linear in windows greater than the

MSE-optimal bandwidths in most groups (as can be seen in **Figure 2** in the main text). However, all estimates were of the same sign as the main estimates in each alternative bandwidth, except for the effect estimate for severe disease outcomes among women, which was close to zero in the main analysis and changed sign from negative to positive with some of the alternative bandwidths (**Tables S4-S5**).

### Supplementary tables

**Table S1.** Estimates of the policy effect on social distancing outcomes from local linear sensitivity analyses varying the bandwidth (BW; i.e., age window) by ±1 and ±2 years from the data-driven MSE-optimal BW. A subtraction indicates a smaller BW (using observations closer to the threshold), whereas an addition gives a larger BW (using more observations farther away from the threshold).

| **Group** | **Bandwidth (BW)** | | | | | All of same sign? |
|---|---|---|---|---|---|---|
| | -2 years | -1 years | MSE-optimal | +1 years | +2 years | |
| *i. Weekly visits to crowded places* | | | | | | |
| All | -.48 (-.98, .03) | -.49 (-.95, -.03) | -.47 (-.89, -.05) | -.41 (-.80, -.01) | -.33 (-.70, .05) | Yes |
| Risk+ | -.33 (-1.08, .42) | -.35 (-1.03, .32) | -.32 (-.95, .31) | -.24 (-.83, .35) | -.14 (-.71, .43) | Yes |
| Risk- | -.6 (-1.26, .05) | -.61 (-1.21, -.02) | -.57 (-1.12, -.02) | -.49 (-1.01, .03) | -.44 (-.93, .05) | Yes |
| Men | -.57 (-1.25, .11) | -.60 (-1.23, .02) | -.56 (-1.14, .02) | -.50 (-1.05, .06) | -.46 (-.99, .07) | Yes |
| Women | -.39 (-1.06, .28) | -.39 (-1.00, .22) | -.35 (-.91, .22) | -.26 (-.79, .27) | -.17 (-.68, .33) | Yes |
| Stockholm | -.84 (-1.88, .20) | -.84 (-1.81, .12) | -.83 (-1.74, .08) | -.80 (-1.66, .07) | -.79 (-1.62, .03) | Yes |
| Rest of Sweden | -.45 (-.99, .09) | -.44 (-.92, .05) | -.38 (-.83, .08) | -.28 (-.71, .15) | -.19 (-.60, .21) | Yes |
| *ii. Number of times gone outside with limited interaction* | | | | | | |
| All | -.44 (-1.22, .35) | -.5 (-1.25, .25) | -.5 (-1.22, .21) | -.49 (-1.18, .2) | -.47 (-1.14, .2) | Yes |
| Risk+ | -.41 (-1.87, 1.05) | -.41 (-1.76, .94) | -.41 (-1.68, .86) | -.38 (-1.59, .83) | -.37 (-1.52, .78) | Yes |
| Risk- | -.51 (-1.48, .47) | -.57 (-1.51, .37) | -.62 (-1.52, .29) | -.63 (-1.51, .24) | -.62 (-1.47, .23) | Yes |
| Men | -.92 (-2.38, .54) | -.95 (-2.25, .36) | -.85 (-2.05, .35) | -.78 (-1.91, .34) | -.71 (-1.77, .35) | Yes |
| Women | -.20 (-1.32, .92) | -.35 (-1.42, .73) | -.42 (-1.45, .62) | -.47 (-1.47, .53) | -.47 (-1.44, .5) | Yes |
| Stockholm | -1.05 (-3.76, 1.65) | -1.35 (-3.77, 1.08) | -1.51 (-3.74, .71) | -1.60 (-3.67, .47) | -1.63 (-3.58, .31) | Yes |
| Rest of Sweden | -.18 (-1.04, .68) | -.23 (-1.05, .60) | -.24 (-1.03, .55) | -.21 (-.98, .55) | -.21 (-.95, .53) | Yes |
| *iii. Weekly visits to healthcare provider(s)* | | | | | | |
| All | .03 (-.02, .08) | .02 (-.02, .06) | .01 (-.02, .05) | .02 (-.02, .06) | .02 (-.01, .06) | Yes |
| Risk+ | .05 (-.02, .13) | .03 (-.03, .1) | .02 (-.03, .08) | .03 (-.03, .09) | .03 (-.04, .09) | Yes |
| Risk- | .01 (-.05, .07) | .01 (-.04, .06) | .01 (-.04, .06) | .02 (-.03, .07) | .03 (-.01, .07) | Yes |
| Men | -.03 (-.08, .02) | -.03 (-.07, .02) | -.02 (-.07, .02) | -.02 (-.06, .03) | -.01 (-.05, .04) | Yes |
| Women | .07 (-.01, .15) | .05 (-.01, .12) | .05 (-.01, .1) | .05 (0.0, .10) | .07 (.01, .12) | Yes |
| Stockholm | .05 (-.04, .13) | .05 (-.03, .13) | .03 (-.04, .1) | .04 (-.03, .10) | .04 (-.02, .1) | Yes |
| Rest of Sweden | .02 (-.03, .08) | .01 (-.03, .06) | .01 (-.04, .05) | .02 (-.03, .06) | .02 (-.02, .06) | Yes |

*Notes:* Estimates reflect bias-corrected absolute effects (where 0 = null effect), with 95% robust confidence intervals from the *rdrobust* package for Stata in paranthases. Risk+ = at least one medical risk factor (see main text), Risk- = no medical risk factors other than old age.

**Table S2.** Estimates of the policy effect on social distancing outcomes from quadratic linear sensitivity analyses varying the bandwidth (BW; age window) by ±1 and ±2 years from the data-driven MSE-optimal BW. A subtraction indicates a smaller BW (using observations closer to the threshold), whereas an addition gives a larger BW (using more observations farther away from the threshold).

| **Group** | **Bandwidth (BW)** | | | | | All of same sign? |
|---|---|---|---|---|---|---|
| | -2 years | -1 years | MSE-optimal | +1 years | +2 years | |
| *i. Weekly visits to crowded places* | | | | | | |
| All | -.36 (-1.27, .55) | -.43 (-1.19, .33) | -.57 (-1.25, .11) | -.70 (-1.32, -.08) | -.69 (-1.27, -.12) | Yes |
| Risk+ | -.21 (-1.41, .98) | -.38 (-1.42, .66) | -.56 (-1.5, .37) | -.64 (-1.5, .23) | -.57 (-1.36, .23) | Yes |
| Risk- | -.56 (-1.53, .41) | -.67 (-1.55, .22) | -.73 (-1.54, .09) | -.74 (-1.5, .02) | -.75 (-1.46, -.04) | Yes |
| Men | -.86 (-1.72, -.01) | -.78 (-1.58, .01) | -.72 (-1.47, .04) | -.64 (-1.36, .08) | -.52 (-1.21, .16) | Yes |
| Women | -.47 (-1.71, .78) | -.59 (-1.66, .49) | -.58 (-1.45, .28) | -.89 (-1.78, .01) | -.85 (-1.68, -.02) | Yes |
| Stockholm | -.85 (-1.97, .26) | -.80 (-1.87, .27) | -.72 (-1.74, .31) | -.66 (-1.65, .33) | -.68 (-1.63, .28) | Yes |
| Rest of Sweden | -.45 (-1.54, .63) | -.53 (-1.41, .35) | -.60 (-1.37, .17) | -.71 (-1.4, -.01) | -.70 (-1.34, -.06) | Yes |
| *ii. Number of times gone outside with limited interaction* | | | | | | |
| All | .06 (-1.16, 1.28) | -.06 (-1.19, 1.07) | -.16 (-1.21, .89) | -.26 (-1.25, .74) | -.32 (-1.26, .62) | No |
| Risk+ | -.37 (-1.86, 1.13) | -.37 (-1.81, 1.06) | -.40 (-1.79, .98) | -.51 (-1.85, .82) | -.62 (-1.91, .67) | Yes |
| Risk- | .67 (-1.05, 2.39) | .40 (-1.16, 1.95) | .21 (-1.22, 1.64) | .00 (-1.34, 1.34) | -.19 (-1.45, 1.07) | No |
| Men | -1.17 (-2.71, .38) | -1.12 (-2.57, .32) | -1.02 (-2.39, .35) | -.93 (-2.24, .37) | -.88 (-2.12, .36) | Yes |
| Women | 1.78 (-.27, 3.84) | 1.43 (-.37, 3.24) | 1.05 (-.58, 2.69) | .75 (-.77, 2.27) | .48 (-.95, 1.91) | Yes |
| Stockholm | -1.32 (-3.95, 1.31) | -1.27 (-3.75, 1.22) | -1.34 (-3.71, 1.02) | -1.45 (-3.72, .81) | -1.70 (-3.87, .48) | Yes |
| Rest of Sweden | .07 (-1.14, 1.28) | .04 (-1.09, 1.17) | -.04 (-1.11, 1.03) | -.10 (-1.11, .92) | -.11 (-1.08, .86) | No |
| *iii. Weekly visits to healthcare provider(s)* | | | | | | |
| All | .05 (-.03, .13) | .04 (-.03, .11) | .01 (-.06, .07) | .01 (-.04, .06) | .01 (-.04, .06) | Yes |
| Risk+ | .03 (-.04, .10) | .03 (-.04, .10) | .03 (-.04, .10) | .03 (-.05, .10) | .02 (-.05, .10) | Yes |
| Risk- | .03 (-.07, .14) | .03 (-.05, .12) | .01 (-.07, .08) | .00 (-.07, .06) | -.02 (-.08, .05) | No |
| Men | -.04 (-.09, .01) | -.03 (-.09, .02) | -.03 (-.08, .02) | -.02 (-.07, .03) | -.01 (-.06, .04) | Yes |
| Women | .05 (-.04, .13) | .01 (-.07, .10) | .02 (-.05, .10) | .04 (-.03, .11) | .05 (-.02, .11) | Yes |
| Stockholm | .04 (-.1, .17) | .07 (-.05, .19) | .05 (-.06, .15) | .03 (-.06, .13) | .02 (-.07, .11) | Yes |
| Rest of Sweden | .03 (-.04, .11) | .00 (-.07, .07) | .00 (-.06, .06) | .00 (-.05, .06) | .00 (-.05, .05) | Yes |

*Notes:* Estimates reflect bias-corrected absolute effects (where 0 = null effect), with 95% robust confidence intervals from the *rdrobust* package for Stata in paranthases. Risk+ = at least one medical risk factor (see main text), Risk- = no medical risk factors other than old age.

**Table S3.** Regression discontinuity estimates for jumps in covariates at the policy threshold using local linear and local quadratic estimation with data-driven (MSE-optimal) bandwidth selection, and covariate-adjusted estimates for the social distancing outcomes.

| Balance check | Local linear | Local quadratic |
|---|---|---|
| *A. Tests for discontinuities in covariates at threshold* | | |
| Women - % | -1.39 (-4.41, 1.62) | -2.86 (-7.27, 1.54) |
| Lives in Stockholm - % | 1.52 (-.88, 3.92) | 1.37 (-1.91, 4.65) |
| Obese (body mass index >= 30) - % | 1.69 (-.68, 4.07) | 2.78 (.08, 5.48) |
| Diabetes - % | -1.89 (-3.53, -.26) | -1.35 (-3.41, .71) |
| Lung disease - % | -1.45 (-3.38, .47) | -1.15 (-3.64, 1.34) |
| Cancer - % | .52 (-.84, 1.88) | 1.23 (-.47, 2.94) |
| Heart disease - % | -.56 (-2.76, 1.65) | -.28 (-2.72, 2.16) |
| Takes immunosuppressants - % | -.40 (-1.80, 1.00) | -1.9 (-4.23, .44) |
| *B. Covariate-adjusted estimates for discontinuity in social distancing outcomes* | | |
| Visits to crowded places | -.50 (-.92, -.07) | -.57 (-1.26, .12) |
| Outside with little interaction | -.50 (-1.20, .21) | -.15 (-1.20, .90) |
| Visits to healthcare provider | .02 (-.02, .05) | .01 (-.05, .07) |

*Notes:* Estimates reflect bias-corrected absolute effects (where 0 = null effect) estimated within MSE-optimal bandwidths, with 95% robust confidence intervals from the *rdrobust* package for Stata in parentheses. We could only perform covariate-adjusted estimation in the social distancing data, since our disease dataset did not contain covariates.

**Table S4.** Estimates of the policy effect on disease outcomes from local linear sensitivity analyses varying the bandwidth (BW; age window) by ±1 and ±2 years from the data-driven MSE-optimal BW. A subtraction indicates a smaller BW (using observations closer to the threshold), whereas an addition gives a larger BW (using more observations farther away from the threshold).

| **Group** | **Bandwidth (BW)** | | | | | All of same sign? |
|---|---|---|---|---|---|---|
| | -2 years | -1 years | MSE-optimal | +1 years | +2 years | |
| *i. Severe cases (dead or hospitalized) per 1,000 population* | | | | | | |
| All | -.91 (-1.85, .03) | -.77 (-1.52, -.03) | -.65 (-1.29, -.01) | -.36 (-.89, .18) | -.51 (-1.09, .07) | Yes |
| Men | -1.25 (-2.42, -.08) | -1.11 (-2.15, -.06) | -.84 (-1.73, .05) | -.65 (-1.49, .19) | -.81 (-1.7, .08) | Yes |
| Women | -.29 (-1.08, .51) | -.18 (-.89, .54) | .03 (-.58, .64) | .11 (-.48, .69) | -.01 (-.63, .61) | No |
| Stockholm | .11 (-1.61, 1.83) | .35 (-1.27, 1.96) | .52 (-1.02, 2.06) | .78 (-.69, 2.26) | .92 (-.5, 2.34) | Yes |
| Rest of Sweden | -1.09 (-2.04, -.14) | -.97 (-1.72, -.21) | -.86 (-1.52, -.21) | -.74 (-1.32, -.15) | -.56 (-1.1, -.02) | Yes |
| *ii. Confirmed cases per 1,000 population* | | | | | | |
| All | NA* | -.34 (-.87, .20) | -.81 (-1.93, .31) | -.60 (-1.28, .08) | -.66 (-1.44, .12) | Yes |
| Men | -1.11 (-2.37, .15) | -1.03 (-2.17, .10) | -.67 (-1.62, .28) | -.46 (-1.38, .46) | -.69 (-1.66, .29) | Yes |
| Women | -.19 (-.86, .47) | -.58 (-1.87, .71) | -.44 (-1.73, .85) | -.25 (-1.08, .57) | -.40 (-1.34, .54) | Yes |
| Stockholm | .40 (-1.47, 2.27) | .63 (-1.13, 2.38) | .83 (-.86, 2.52) | 1.29 (-.30, 2.89) | 1.65 (.11, 3.19) | Yes |
| Rest of Sweden | NA* | -.48 (-1.04, .07) | -.78 (-2.02, .46) | -.87 (-1.69, -.05) | -.85 (-1.55, -.14) | Yes |

*Notes:* Estimates reflect bias-corrected absolute effects (where 0 = null effect), with 95% robust confidence intervals from the *rdrobust* package for Stata in paranthases. *Insufficient observations to run a local regression (MSE-optimal bandwidth too close to threshold).

**Table S5.** Estimates of the policy effect on disease outcomes from local quadratic sensitivity analyses varying the bandwidth (BW; age window) by ±1 and ±2 years from the data-driven MSE-optimal BW. A subtraction indicates a smaller BW (using observations closer to the threshold), whereas an addition gives a larger BW (using more observations farther away from the threshold).

| **Group** | **Bandwidth (BW)** | | | | | All of same sign? |
|---|---|---|---|---|---|---|
| | -2 years | -1 years | MSE-optimal | +1 years | +2 years | |
| *i. Severe cases (dead or hospitalized) per 1,000 population* | | | | | | |
| All | -1.06 (-2.42, .3) | -1.05 (-2.16, .06) | -.99 (-1.94, -.03) | -.94 (-1.78, -.09) | -.89 (-1.66, -.12) | Yes |
| Men | -1.07 (-3.29, 1.16) | -1.33 (-3.14, .49) | -1.47 (-2.86, -.08) | -1.28 (-2.68, .13) | -1.23 (-2.52, .05) | Yes |
| Women | -.71 (-2.17, .76) | -.66 (-1.87, .56) | -.32 (-1.32, .67) | -.52 (-1.48, .43) | -.52 (-1.40, .36) | Yes |
| Stockholm | -1.14 (-3.95, 1.67) | -.80 (-3.32, 1.72) | -.44 (-2.74, 1.86) | -.68 (-2.83, 1.48) | -.47 (-2.51, 1.56) | Yes |
| Rest of Sweden | -1.24 (-2.66, .17) | -1.20 (-2.33, -.06) | -1.24 (-2.2, -.29) | -1.20 (-2.06, -.34) | -1.12 (-1.90, -.34) | Yes |
| *ii. Confirmed cases per 1,000 population* | | | | | | |
| All | -.85 (-2.85, 1.15) | -.78 (-2.24, .68) | -.73 (-2.02, .56) | -1.11 (-2.16, -.07) | -1.32 (-2.27, -.38) | Yes |
| Men | -.59 (-3.32, 2.15) | -.72 (-2.9, 1.46) | -1.46 (-3.11, .20) | -.94 (-2.57, .70) | -1.14 (-2.62, .34) | Yes |
| Women | -.82 (-3.42, 1.78) | -.97 (-2.83, .89) | -.43 (-1.97, 1.12) | -1.33 (-2.66, .00) | -1.62 (-2.82, -.41) | Yes |
| Stockholm | -2.23 (-6.2, 1.74) | -1.64 (-4.88, 1.59) | -.29 (-3.34, 2.76) | -.62 (-3.16, 1.92) | -.72 (-3.06, 1.62) | Yes |
| Rest of Sweden | -.43 (-2.51, 1.64) | -.60 (-2.09, .89) | -.81 (-2.13, .51) | -1.37 (-2.45, -.3) | -1.70 (-2.67, -.73) | Yes |

*Notes:* Estimates reflect bias-corrected absolute effects (where 0 = null effect), with 95% robust confidence intervals from the *rdrobust* package for Stata in paranthases. *Insufficient observations to run a local regression (MSE-optimal bandwidth too close to threshold).

### Supplementary figures

##### List of Supplementary figures

**Figure S1.** Regression discontinuity plots for the impact of the age-specific isolation recommendations on social distancing behaviors at the 70-year-threshold with local linear estimates in mean-squared-error-optimal bandwidths around the threshold for subgroups based on medical risk factors, sex and area (Stockholm county, rest of Sweden), for three social distancing measures: A) average weekly visits to crowded places, B) average weekly outdoor episodes with no or limited interaction, and C) average weekly visits to healthcare providers. ..... 14

**Figure S2.** Regression discontinuity plots for the impact of the age-specific isolation recommendations on social distancing behaviors at the 70-year-threshold with local quadratic estimates in mean-squared-error-optimal bandwidths around the threshold for subgroups based on medical risk factors, sex and area (Stockholm county, rest of Sweden), for three social distancing measures: A) average weekly visits to crowded places, B) average weekly outdoor episodes with no or limited interaction, and C) average weekly visits to healthcare providers. ..... 15

**Figure S3**. Regression discontinuity plots for visits to crowded places in detailed subgroups by risk factor status and sex. ..... 16

**Figure S4.** Regression discontinuity plots for the impact of the age-specific isolation recommendations on COVID-19 disease incidence per 1,000 population at the 70-year-threshold with local linear estimates in mean-squared-error-optimal bandwidths around the threshold in subgroups by sex and area (Stockholm county, rest of Sweden). A) Severe cases (hospitalized or dead), B) all confirmed cases. The incidence is presented on a logarithm scale to enable better visualization of the regions around the 70-year-threshold. ..... 17

**Figure S5.** Regression discontinuity plots for the impact of the age-specific isolation recommendations on COVID-19 disease incidence per 1,000 population at the 70-year-threshold with local quadratic estimates in mean-squared-error-optimal bandwidths around the threshold in subgroups by sex and area (Stockholm county, rest of Sweden). A) Severe cases (hospitalized or dead), B) all confirmed cases. The incidence is presented on a logarithm scale to enable better visualization of the regions around the 70-year-threshold. ..... 18

**Figure S6.** Histogram of the forcing variable age showing the frequency of observations to check for evidence of sorting of observations around the 70-year-threshold (policy threshold is indicated with a vertical line). The figure does not show evidence of sorting, as the number of observations develops smoothly across the threshold. ..... 19

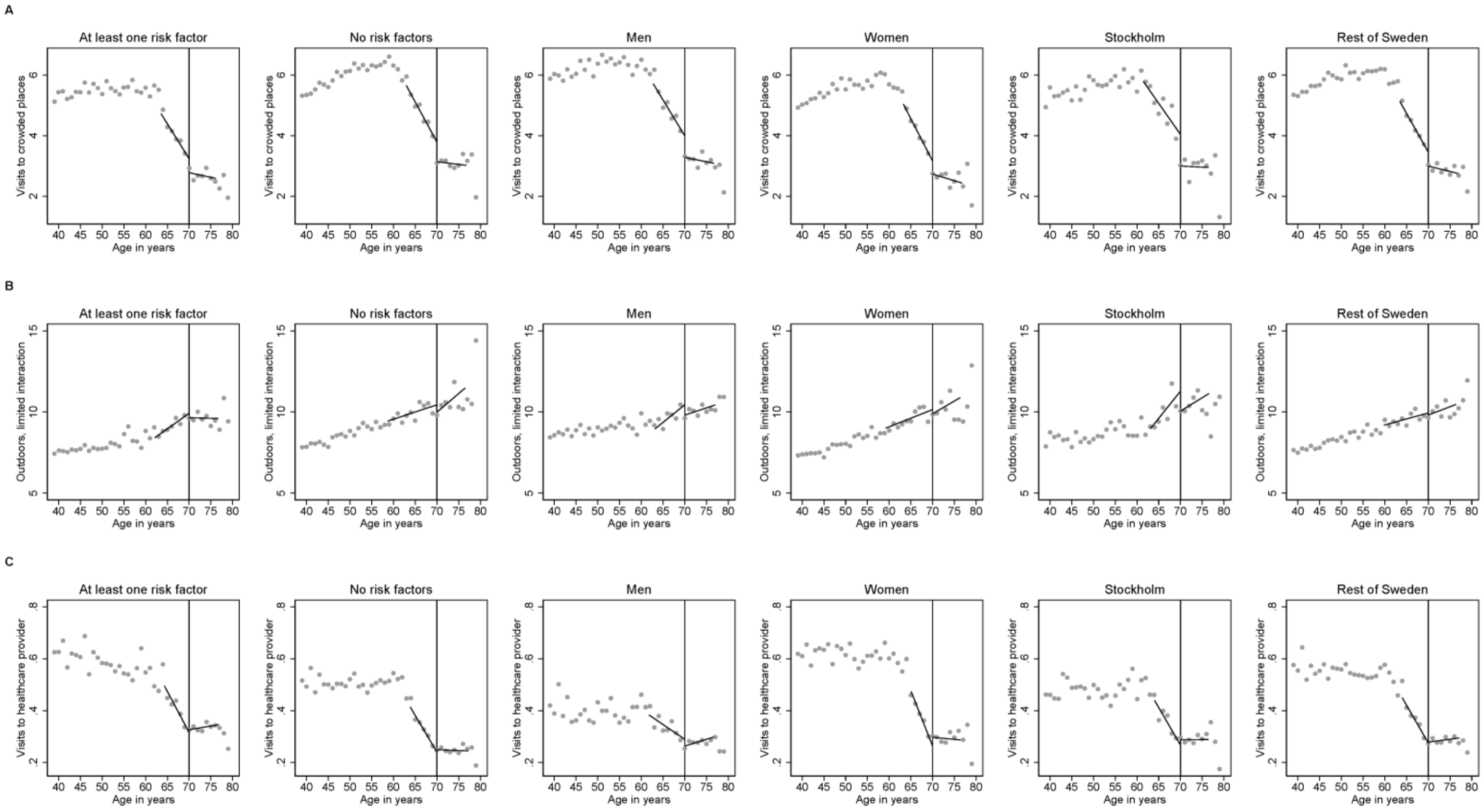

**Figure S1.** Regression discontinuity plots for the impact of the age-specific isolation recommendations on social distancing behaviors at the 70-year-threshold with local linear estimates in mean-squared-error-optimal bandwidths around the threshold for subgroups based on medical risk factors, sex and area (Stockholm county, rest of Sweden), for three social distancing measures: A) average weekly visits to crowded places, B) average weekly outdoor episodes with no or limited interaction, and C) average weekly visits to healthcare providers.

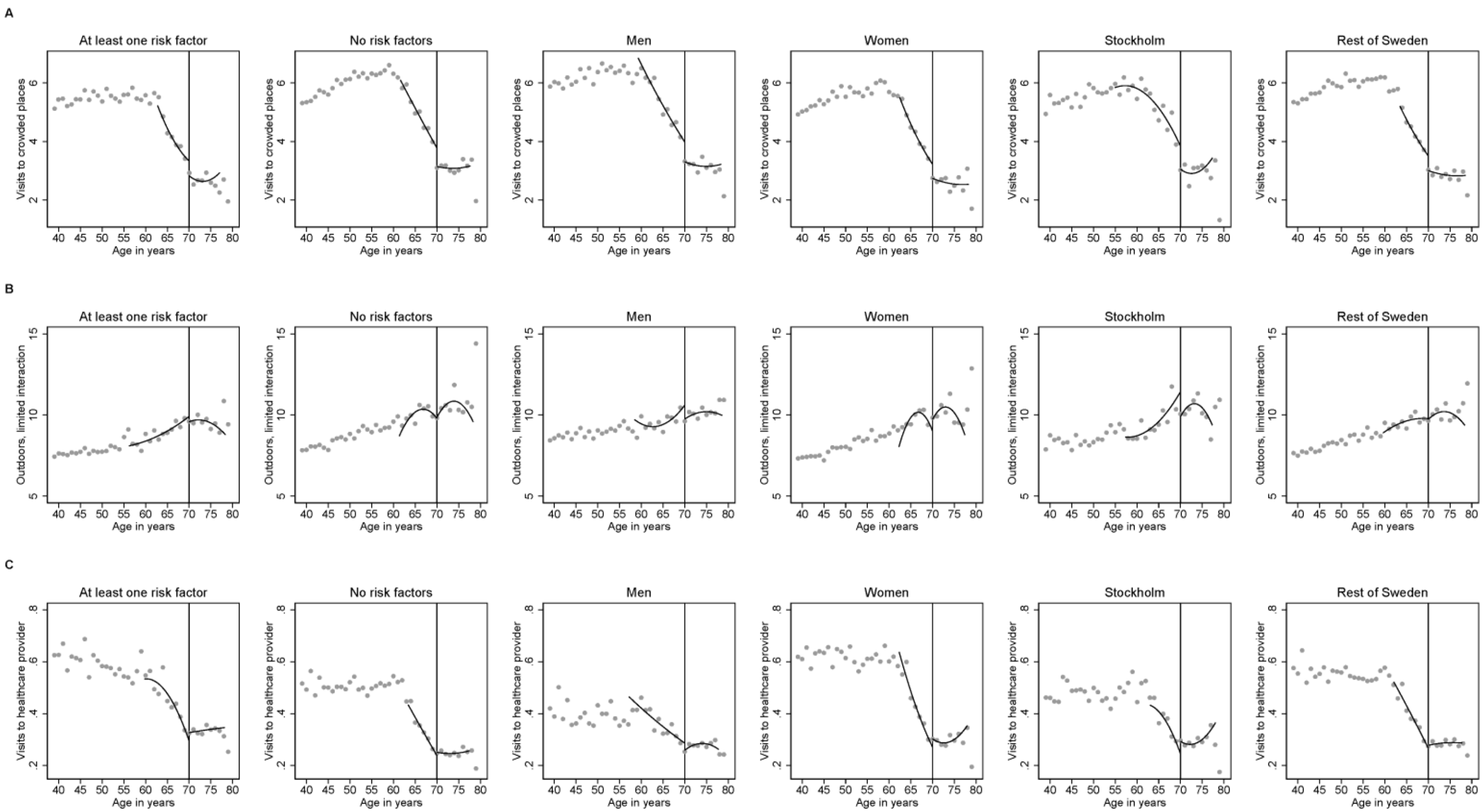

**Figure S2.** Regression discontinuity plots for the impact of the age-specific isolation recommendations on social distancing behaviors at the 70-year-threshold with local quadratic estimates in mean-squared-error-optimal bandwidths around the threshold for subgroups based on medical risk factors, sex and area (Stockholm county, rest of Sweden), for three social distancing measures: A) average weekly visits to crowded places, B) average weekly outdoor episodes with no or limited interaction, and C) average weekly visits to healthcare providers.

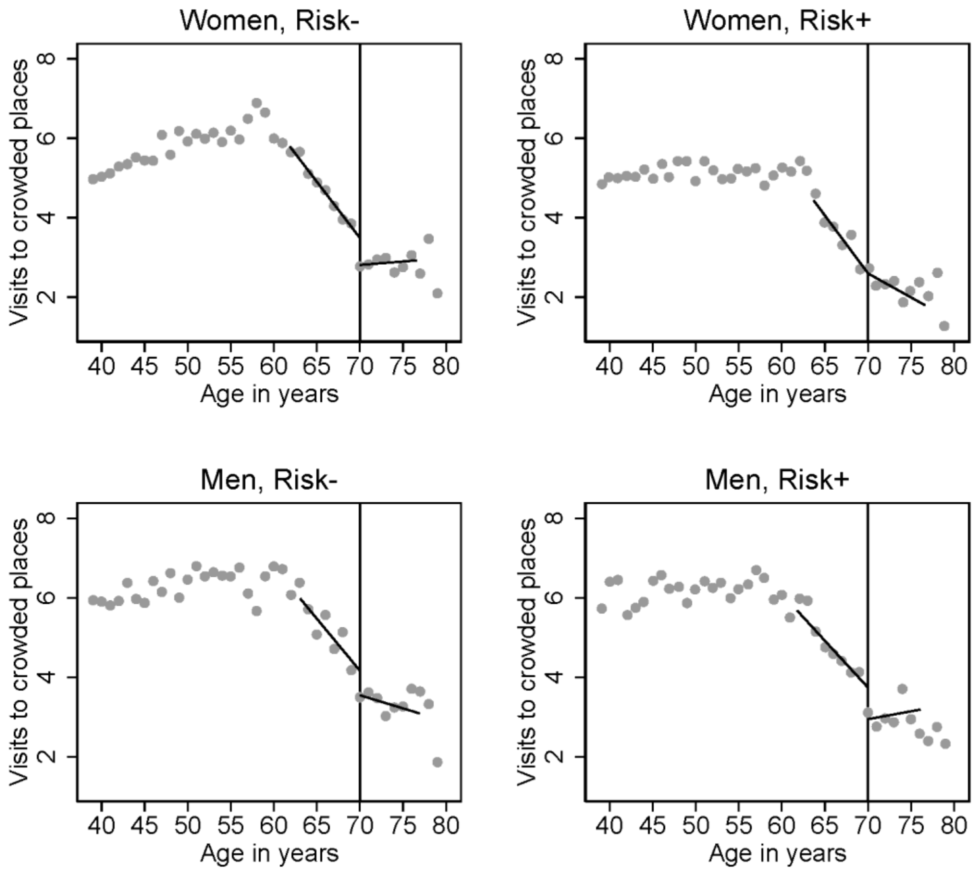

**Figure S3**. Regression discontinuity plots for visits to crowded places in detailed subgroups by risk factor status and sex.

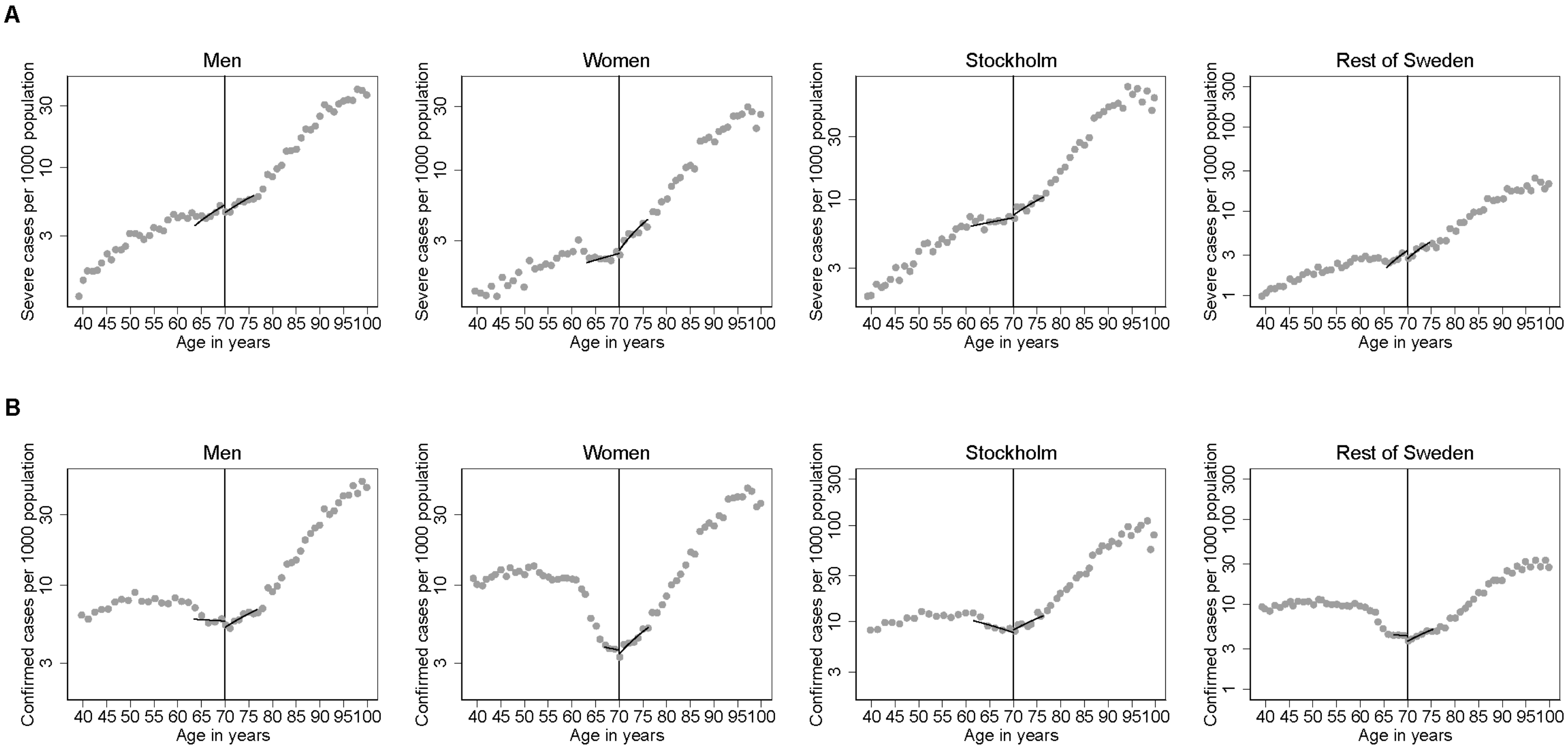

**Figure S4.** Regression discontinuity plots for the impact of the age-specific isolation recommendations on COVID-19 disease incidence per 1,000 population at the 70-year-threshold with local linear estimates in mean-squared-error-optimal bandwidths around the threshold in subgroups by sex and area (Stockholm county, rest of Sweden). A) Severe cases (hospitalized or dead), B) all confirmed cases. The incidence is presented on a logarithm scale to enable better visualization of the regions around the 70-year-threshold.

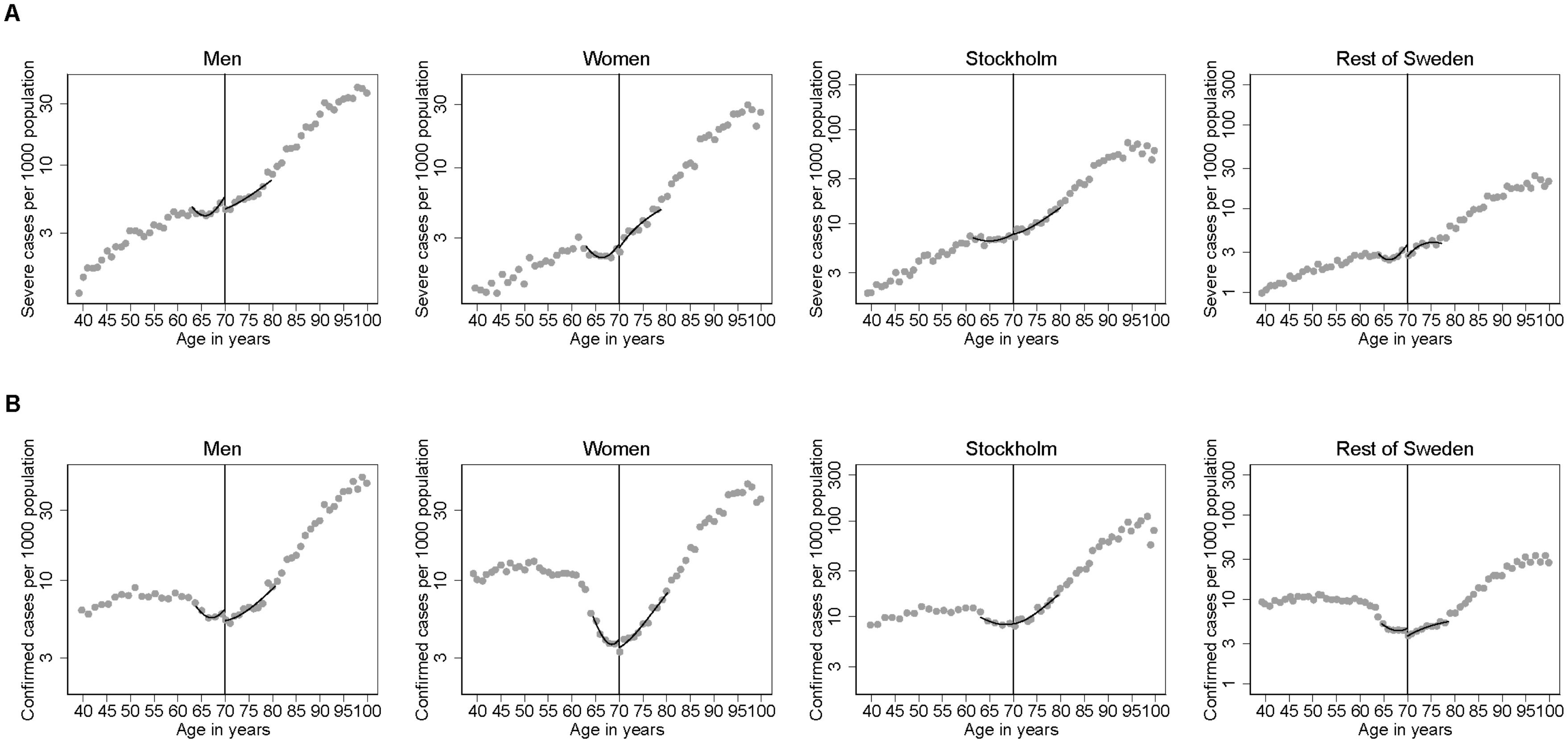

**Figure S5.** Regression discontinuity plots for the impact of the age-specific isolation recommendations on COVID-19 disease incidence per 1,000 population at the 70-year-threshold with local quadratic estimates in mean-squared-error-optimal bandwidths around the threshold in subgroups by sex and area (Stockholm county, rest of Sweden). A) Severe cases (hospitalized or dead), B) all confirmed cases. The incidence is presented on a logarithm scale to enable better visualization of the regions around the 70-year-threshold.

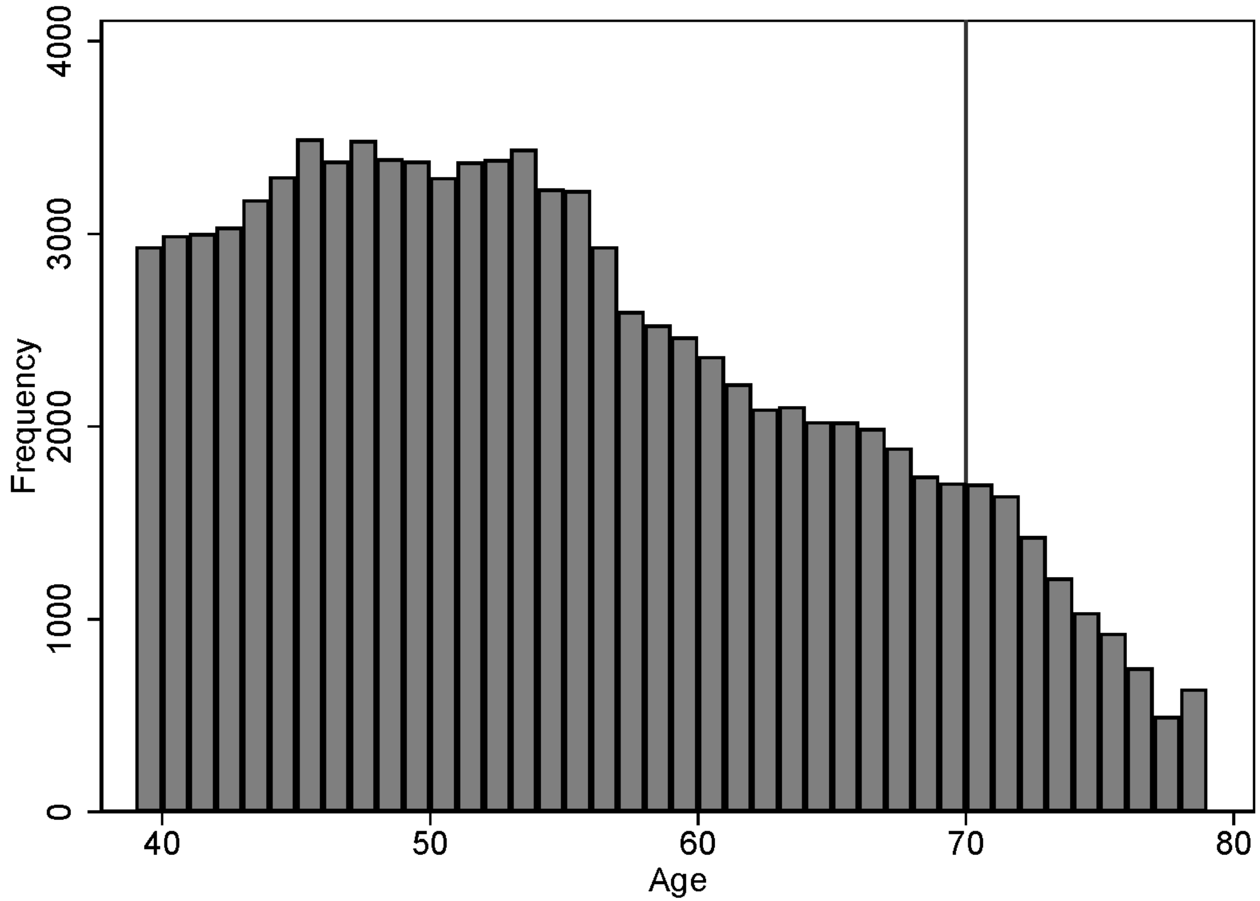

**Figure S6.** Histogram of the forcing variable age showing the frequency of observations to check for evidence of sorting of observations around the 70-year-threshold (policy threshold is indicated with a vertical line). The figure does not show evidence of sorting, as the number of observations develops smoothly across the threshold.
